# Characterizing US spatial connectivity: implications for geographical disease dynamics and metapopulation modeling

**DOI:** 10.1101/2023.11.22.23298916

**Authors:** Giulia Pullano, Lucila G. Alvarez-Zuzek, Vittoria Colizza, Shweta Bansal

## Abstract

**Background:** Human mobility is expected to be a critical factor in the geographic diffusion of infectious diseases, and this assumption led to the implementation of social distancing policies during the early fight against the COVID-19 emergency in the United States. Yet, because of substantial data gaps in the past, what still eludes our understanding are the following questions: 1) How does mobility contribute to the spread of infection within the United States at local, regional, and national scales? 2) How do seasonality and shifts in behavior affect mobility over time? 3) At what geographic level is mobility homogeneous across the United States? Addressing these questions is critical to developing accurate transmission models, predicting the spatial propagation of disease across scales, and understanding the optimal geographical and temporal scale for the implementation of control policies.

**Methods:** We address this problem using high-resolution human mobility data measured via mobile app usage. We compute the daily connectivity network between US counties to understand the spatial clustering and temporal stability of mobility patterns. We then integrate our mobility data into a spatially explicit transmission model to reproduce the national invasion of the first wave of SARS-CoV-2 in the US, and characterize the impact of the spatio-temporal scale of mobility data on disease predictions.

**Findings:** Temporally, we observe that intercounty connectivity is annually stable, and was unperturbed by mobility restrictions during the early phase of the COVID-19 pandemic, despite significant changes in overall activity. Spatially, we identify 104 geographic clusters of US counties that are highly connected by mobility within the cluster and more sparsely connected to counties outside the cluster. Together, these results suggest that intercounty connectivity in the US is relatively static across time and is highly connected at the sub-state level. We find that the stability in temporal patterns allows static mobility data to effectively capture infection dynamics. On the other hand, spatial uniformity at the sub-state (cluster)-scale does not capture spatial dynamics; instead, mobility data at the county-scale is necessary to better predict spatial disease diffusion.

**Interpretation:** Our work demonstrates that intercounty mobility was negligibly affected outside the lockdown period of Spring 2020, explaining the broad spatial distribution of COVID-19 outbreaks in the US during the early phase of the pandemic. Such geographically dispersed outbreaks place a significant strain on national public health resources and necessitate complex metapopulation modeling approaches for predicting disease dynamics and control design. We thus inform the design of such metapopulation models to balance high disease predictability with low data requirements.

## Introduction

Human mobility plays a crucial role in the spread of respiratory diseases [17, 41]. The combination of regional travel and local commuting represents the spatial connectivity between locations, serving as the main driver in the geographic diffusion of infectious diseases. Characterizing the spatial dynamics of pathogen transmission is, therefore, intricately tied to unraveling human mobility patterns. Such a task has proven to be challenging due to the inherent complexity and privacy-related limitations on collecting mobility data [7]. Over the past few decades, researchers have extensively relied on mobility data obtained from census records, surveys, transportation statistics, commuting data, and international air traffic data. Such datasets have widely contributed to a better understanding of human mobility patterns and their impact on the epidemic spread [1, 6, 3, 11, 39, 2, 33], but can be limited in their resolution or scale. More recently, this gap has been filled by the use of mobile phone data [10, 14], primarily based on phone records, but no such data has been available in the United States.

The global health crisis triggered by COVID-19 has underscored the critical need for swift access to mobility to help mitigate the spread of the virus. The urgency of the situation prompted an unprecedented sharing of data by private companies worldwide, through legally and ethically compliant agreements. This data was based on mobile location-based app usage and thus provided incomparable access to high-resolution, large-scale, and near-real-time mobility data and has expanded human mobility science [37], and computational epidemiology [19, 18]. The availability of this data has especially represented a shift in US public health and it has been used to inform epidemic models and reveal the impact of mitigation strategies on behavior[27, 38, 35, 32, 20, 26, 15, 25]. While the association between mobility patterns and COVID-19 transmission in the USA has been extensively studied, no studies have been devoted to assessing when the underlying mobility network needs to be embedded into models to characterize epidemic spread.

Moreover, the effects of control measures on human mobility at mesoscale (i.e., intermediate or regional level of geographical granularity) and long-range (i.e., entire countries, continents), as well as the most suitable geographical and temporal granularity for implementing these measures, still lack clarity. This gap in understanding the characteristic spatio-temporal scale of mobility not only limits target control policies but also our ability to model transmission dynamics effectively. To date, mobility data have been integrated into epidemic models without due consideration for the optimal geographical (e.g. municipalities, regions, states) and temporal resolution (e.g., day, week, month) required to accurately capture epidemic spread. The level of granularity used in these models has consistently been dictated by a priori assessments from data providers [28, 34].

To address these gaps, we aim to characterize the spatiotemporal characteristic scale of human mobility in the United States, for the periods before and after the pandemic emergency by using mobile phone data. Furthermore, to assess if mobility was relevant in the spread of COVID-19 and which mobility scale drove the invasion, we integrate mobility data into spatially-explicit transmission models to reproduce the national invasion of the first wave of SARS-CoV-2 in the US. More specifically, we observe the daily intercounty connectivity in the US in time and space. To assess the role of mobility, we evaluate how the predictability of the model depends on the underlying mobility data scale, and how model predictive power is impacted by not accounting for fine-grain mobility.

## Methods

Our study has two objectives: i) to characterize the temporal and spatial scale of variation in the connectivity between U.S. counties influenced by human mobility and ii) to inform metapopulation models with mobility data at different scales, thereby informing the scale of data required for epidemic predictability. To achieve this, we analyze human mobility data measured via mobile app usage across spatial and temporal scales. We then integrate these mobility data into an epidemic model parameterized with national COVID-19 public health data to evaluate the performance of the model at different scales of mobility to predict epidemic spatial invasion.

### Characterizing intercounty connectivity with mobility data

We access data from SafeGraph [45] (now called Advan Patterns), which collects and shares mobility data based on location-based mobile app usage. In particular, we use the daily Social Distancing dataset provided by Safegraph (see Supplementary Information for dataset selection), and use information on the number of mobile devices with a home in an origin census block group that visit a given destination census block group or staying in the origin one for at least a minute. Data span the period from January 2019 to April 2021 on a daily timescale. We aggregate the data to the US county level to capture a common geographic scale for disease surveillance and public health decision-making. To address the spatial and temporal heterogeneity in the observed devices obs_*i*_ within each county *i* (Figure S1), we developed a correction factor:

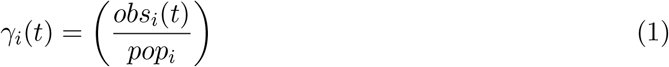

where *pop*_*i*_ is the population in the county *i*. To reduce sampling biases, we exclude the bottom 25% of counties by population size, running all analyses on 2327 geographical counties within the continental US of a population size greater than 11,000.

After rescaling for the correction factor, to quantify intercounty connectivity monthly, we normalize the number of visits between an origin and destination county by the daily total number of visits originating in the origin county. We then compute the average daily visits each month from Jan 2019 to March 2021 for all pairs of counties. In Figure 1A is shown for illustrative proposes the spatial connectivity network computed in March 2020. We therefore obtain a time-evolving connectivity network between US counties, in which the links represent the daily coupling probabilities *p*_*ij*_ between any pair *i* and *j* of US counties. *p*_*ij*_ is normalized as follows: Σ_*i*≠*j*_ *p*_*ij*_ +*p*_*ii*_ = 1. A comparison between the monthly and daily dataset provided by SafeGraph is reported in Figure S2.

**Figure 1:**
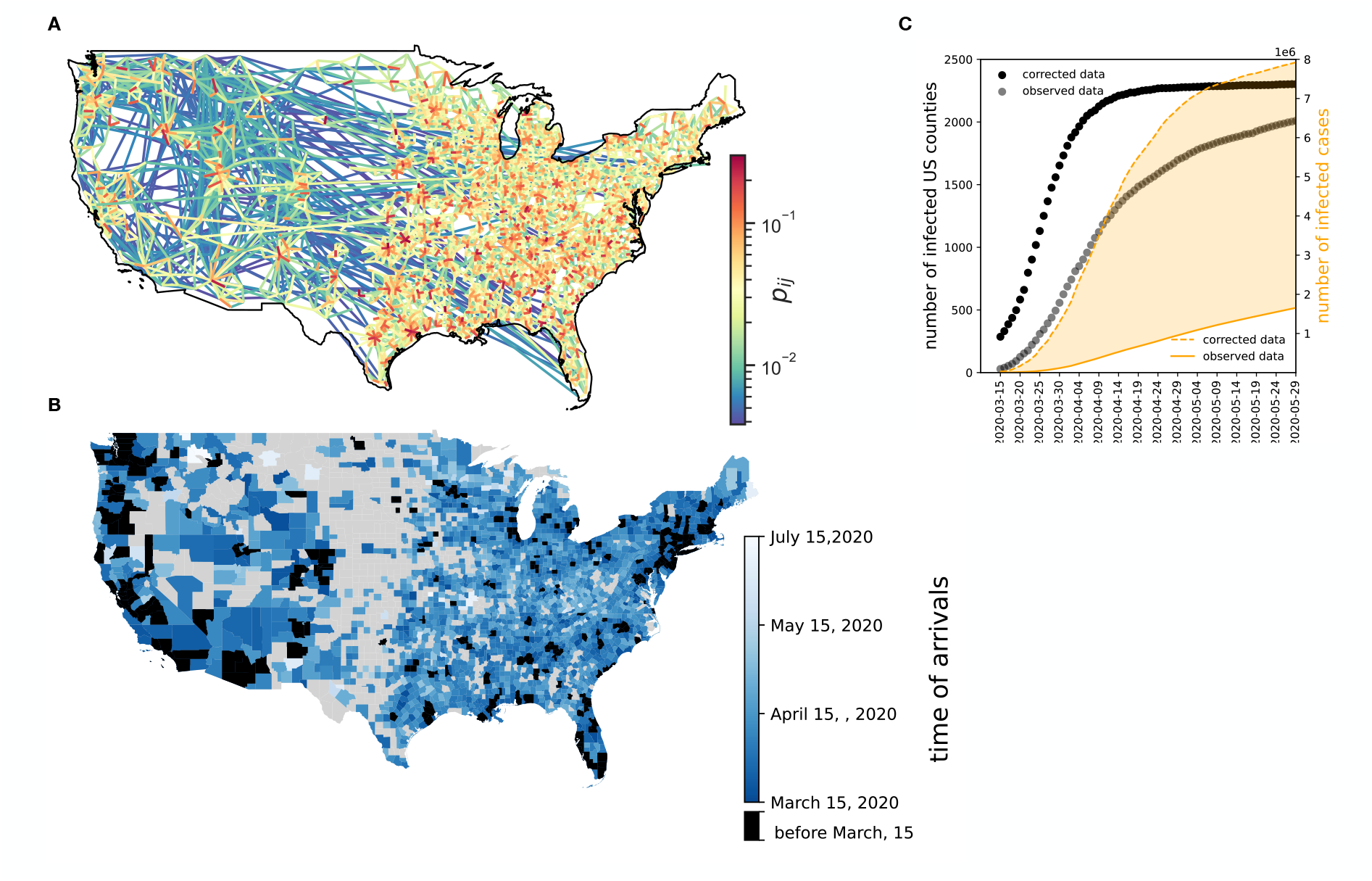
Data sources and epidemic context. (A) Mobility data. Figure shows the spatial connectivity network between US counties on March, 2020. Only links with top 1% of coupling probability are shown. (B-C) Public health data. (B) Color coded map shows the corrected time of arrival by county i.e. the time when at least 10 infected cases are reported. Black colored counties are counties that have been infected before March, 15. (C) Black dots show the corrected number of infected counties over time. A county is considered infected if it reports at least 10 confirmed COVID-19 cases. Grey dots show the observed number of infected counties over time. A county is considered infected if it reports at least 10 confirmed COVID-19 cases.Orange solid line shows the daily number of new confirmed cases nationally, while the orange dotted line shows the real number of cases accounting for underreporting.

### Early phase of COVID-19 in the US

The initial confirmed case of COVID-19 in the United States was reported in Washington state on January 21, and within a few weeks, local transmission was established. Guidelines advocating for social distancing and the avoidance of gatherings were released on March 16. During the early stages of the COVID-19 pandemic, while most European countries implemented national restrictions, lockdowns and stay-at-home orders in the United States -like in Chile - were implemented locally and at diverse times. The peak of lockdowns was in April 2020, when more than 40 states had issued some form of stay-at-home or shelter-in-place order [16]. However, the spatial spread of COVID-19 was not contained, and at the end of June 2020, most US counties reported COVID-19 cases. Facing a rebound of cases in the fall of 2020, social distancing was recommended (but not mandated) to maintain epidemic activity at low levels.

COVID disease incidence data was derived from CDC data [**empty citation**]. In this work, we use COVID-19-related daily new reported cases and the time of arrival in any US county, i.e. the day when at least 10 cases have been reported in that area. daily new reported cases and time of arrivals have been corrected by county accounting for undereporting. We estimate the level of ascertainment using reported data on COVID-19 cases and fatalities globally, as described in [**russell“reconstructing“2020**]. (Figure 1 B-C).

### Describing temporal and spatial variability in the mobility network

We examine the monthly network structure to evaluate the temporal dynamics of mobility patterns. We quantify the degree by county i.e. the number of connections (edges) the counties in the network have to other counties. We also define link persistence as the probability that links between counties that exist with non-zero mobility during the month of 2019 remain present in the same month during 2020 and 2021.

We also fitted a gravity model to the intercounty connectivity network for each month. For technical details and model performance, see Supplementary Information (SI).

To detect the different geographical communities generated by human mobility patterns, we performed a community detection analysis using the stochastic InfoMap algorithm [13]. We aim to identify regions where within movements occur more frequently compared to movements to other regions. To account for stochasticity, we use a bootstrap resampling method (see SI for details).

The classification of urban and rural areas was determined based on the NCHS Urban-Rural Classification Scheme for counties [31].

All network analyses were done using Python’s `networkX` library, and the gravity model fitting was done using the Python `scikit-mobility` library.

### Incorporating human mobility into infectious disease models

We used a stochastic non-Markovian transmission model with a metapopulation structure at the US county level [44]. In each county, the model accounted for disease transmission proportional to (i) infected residents not moving (ii) infected visitors coming from other counties and (iii) returning residents previously infected in other counties. The resulting force of infection in the county *i* is defined as follows:

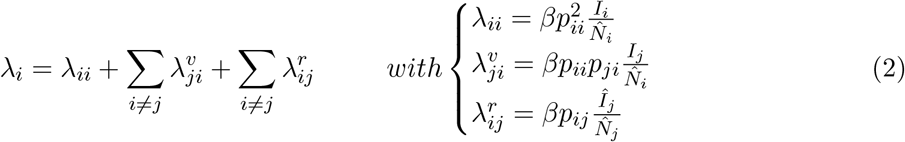

where *p*_*ij*_ is the coupling probability between patches *i* and *j* extracted from the intercounty connectivity network. The effective population, and effective number of infections, are respectively defined as follows:

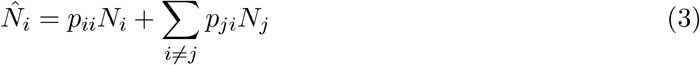

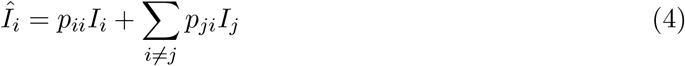

We considered SEIR (Susceptible - Exposed - Infectious - Recovered) epidemic dynamics specific to COVID-19. Epidemics parameters are informed by [29].

The detailed mathematical framework, model calibration and implementation details can be found in the Supplement.

### Inference framework and Goodness of fit

To effectively calibrate the epidemic pathway, we utilize national-level data on the count of counties with a minimum of 10 reported infected cases over time (see Figure 1C). The calibration process was conducted within the time frame spanning March 14, 2020, to Sept 15, 2020, a period during which all counties had reported instances of infection. During this process, we derive the parameter estimates *β*_*pre−LD*_ for the time interval from March 15 to March 31, and *β*_*post−LD*_ for the period from March 31 to May 15, 2020. Inference is based on maximum likelihood, assuming a Poisson distribution to model the reported number of infected cases over time.

To assess model performance, we computed the goodness of fit to compare the estimated invasion probability *p*_*i,inv*_(*t*) with the observed early phase COVID-19 spatial invasion. *p*_*i,inv*_(*t*) denotes the likelihood for a county *i* in a day *t* to report at least 10 infected cases in the simulation [9]. The goodness of fit is defined as follows:

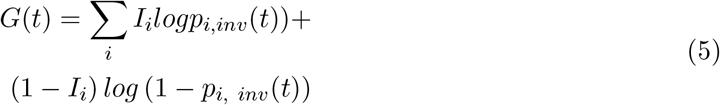

*I*_*i*_ = 1 if the county *i* have been at least 10 infected cases at the day *t*, and *I*_*i*_ = 0 otherwise.

### Comparing models across geographical scales

In designing the metapopulation structure across various spatial scales, we maintain the spatial scale of the model at the county level. This approach enables us to make meaningful comparisons of the results across different scales. To specify a metapopulation model at a spatial scale *R* (e.g., cluster, state), we randomize the mobility links that occur among all counties within a patch at scale *R*, preserving the number of links. We then set the coupling probabilities for the connected counties within each patch *R* equal to the average coupling probability of the links within the patch, as follows:

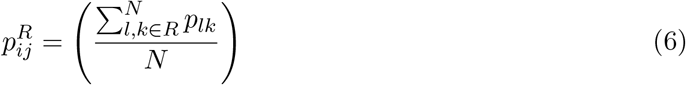

with *i, j* ∈ *R*, while the coupling probabilities across patches are not changed.

## Results

Human mobility is expected to be a pivotal driver in infectious disease transmission. Understanding the impact of mobility on infection spread at local, regional, and national scales is therefore imperative for precision in transmission models, predicting disease spread, and optimizing targeted control strategies. By analyzing US county-level spatial connectivity using mobile phone data, we assess the temporal and geographical variability of human mobility and identify the geographical scale that drove the early phase of COVID-19 spatial invasion. As we address public health questions, we reveal the characteristic scale to design metapopulation modeling.

### Temporal stability of the intercounty connectivity network

Limited changes in mobility are observed from January 2019 to March 2021, except for a significant impact localized to April 2020 (Figure 2A). This notable transition coincided with the implementation of lockdown measures, causing a nationwide decline in mobility from roughly 45 million daily visits to about 25 million visits post-lockdown enforcement. The mobility shock extended throughout the month, encompassing a transitional period (Figure 2A-B). Analyzing the temporal evolution of the intercounty connectivity network, we discovered a consistent seasonal pattern in the degree distribution and the persistence of mobility connections. Local variations were observed, only in April 2020, with a 23% reduction in degree and a 26% reduction in link persistence, respectively. Surprisingly, no variation was observed in November 2020, despite the strong recommendations for social distancing ahead of the winter surge of SARS-CoV-2. The reduction in Rural-Urban connections is particularly pronounced, with a 25% decrease compared to the pre-reduction value in February 2020. This decrease stabilized in May and beyond. Notably, Urban-Urban connections exhibited greater resilience over time when compared to connections involving rural areas. Moreover, while coupling probabilities stay consistent over the study period (Figure 1C), the likelihood of staying in the home location exhibited larger variability (See SI).

**Figure 2:**
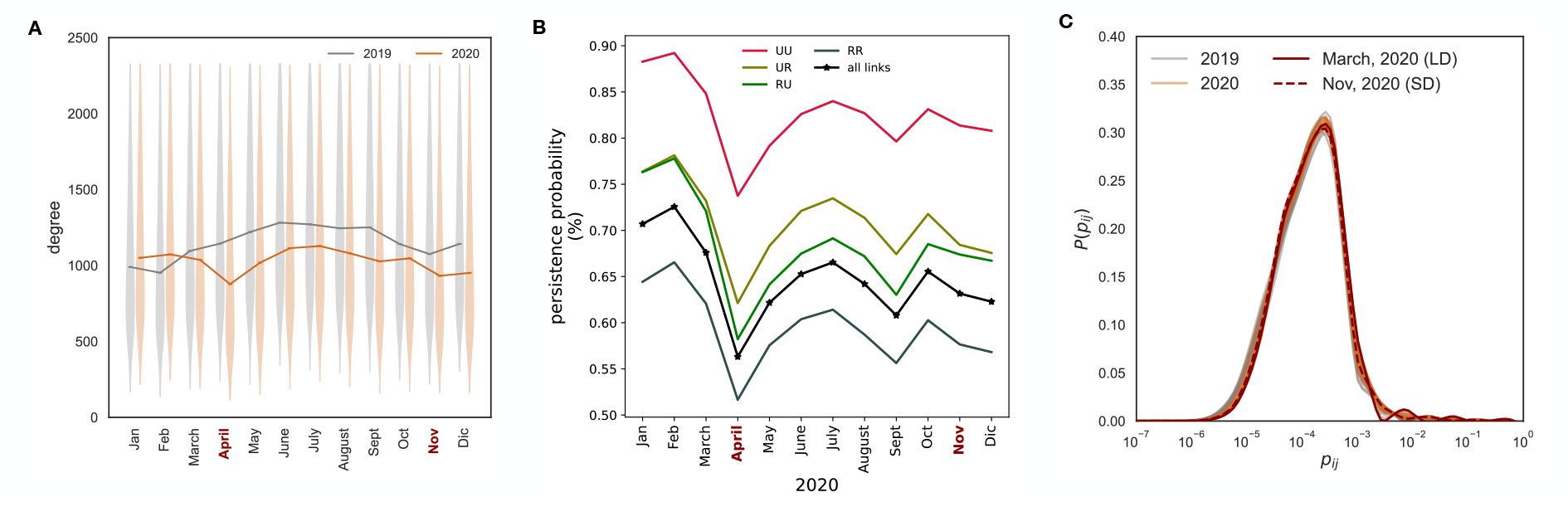
Temporal stability in intercounty mobility. (A) The monthly degree distribution of the inter- county network show significant variability but are consistent across months. (B) The persistence probability of links is illustrated, denoting the likelihood that a connection existing in 2019 remains present in 2020 and 2021. The plot provides a breakdown for different link types: urban-urban (UU), urban-rural (UR), rural-urban (RU), and rural-rural (RR) links. (C) Distribution of coupling probabilities in the connectivity network by month. We highlighted in darkred i) April, 2020 (LD) that represents the peak time of number of US states in lockdown, and November, 2020 (SD) that represents the period when social distancing recommendations were in place.

In spite of sporadic extreme events leading to local variability, the intercounty connectivity network demonstrated temporal stability and exhibited a high level of predictability through a gravity fit model as shown in Figure S7-9. Indeed, the Spearman coefficient between the original and modeled intercounty connectivity network remains constant over time, averaging 0.55.

### Spatial stability of the intercounty connectivity network

To identify the geographic scale at which mobility is highly connected, we detect clusters of counties that are more connected via mobility within the cluster than outside the clusters, we use a network community detection algorithm. Our hypothesis was that this partitioning of the US would be at a geographic scale larger than 3143 US counties but smaller than 50 US states or 10 HHS regions. Indeed, we find that based on human mobility, the US can be partitioned into around 100 regions that split most US states into multiple clusters (Figure 3). We also find that these clusters are highly spatially contiguous and respect state boundaries (with a similarity measured by normalized mutual information (NMI) as 0.82). Furthermore, these regions demonstrate stability over time (NMI = 0.95) despite the perturbations of the early phases of the COVID pandemic (Figure 3B and in Figure S9-10). Thus, we identify a persistent geographic partitioning of the US in which clusters are more connected within than between, and hypothesize that the relevance of mobility to the spatial diffusion of infectious diseases occurs at a mesoscale.

**Figure 3:**
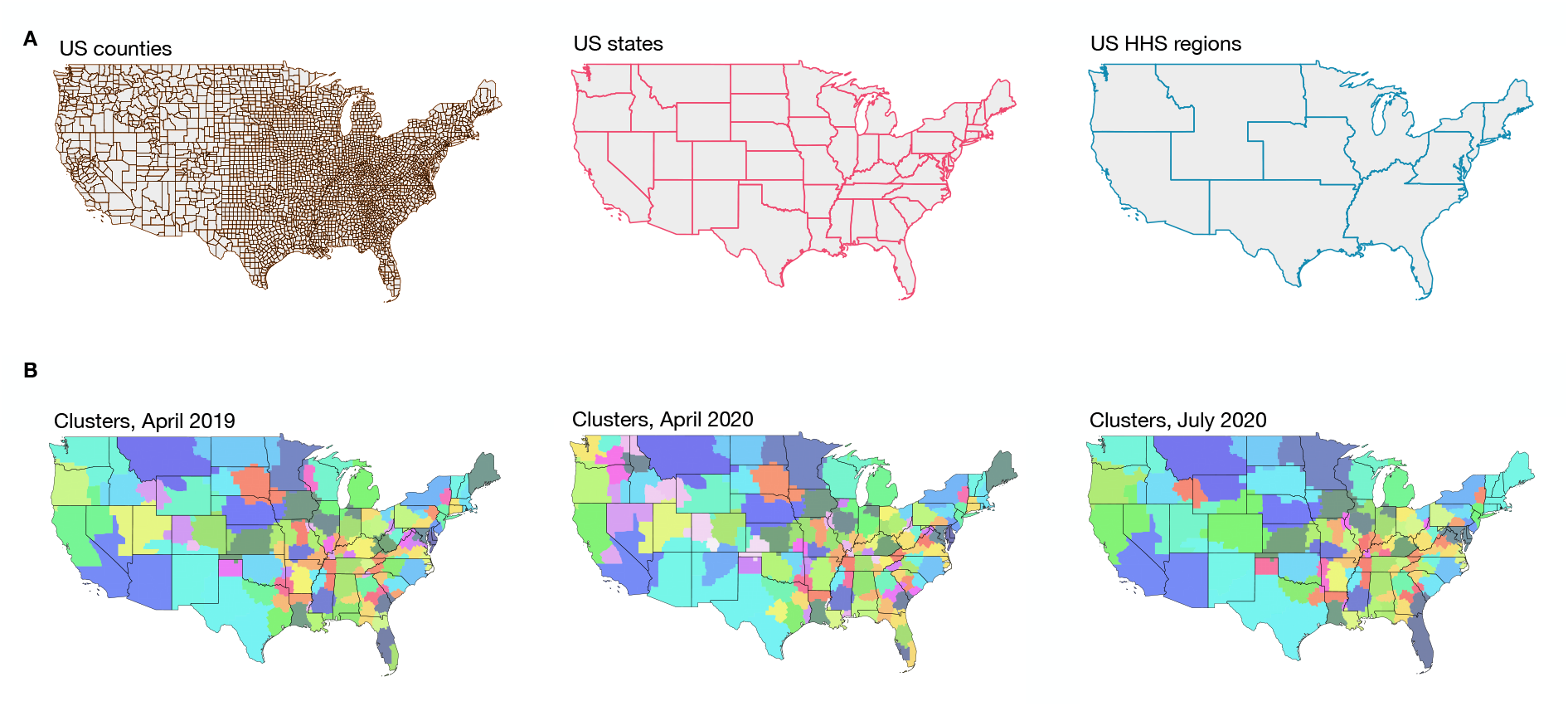
Analysis of Spatial Stability. (A) Geographical subdivisions at the county and state levels within the US, as well as the division into HHS (Health and Human Services) regions, used for health administration purposes. (B) We partition the intercounty mobility network so that each cluster of counties is more strongly connected to each other via mobility than to counties in other clusters. We find highly consistent partioning based on mobility networks from April 2019, April 2020 (during the mandated lockdown period of the early COVID pandemic) and July 2020 (after the lockdown period of the early COVID pandemic). Clusters are colored to delineate cluster boundaries and do not represent any other information. Counties colored in gray have populations of fewer than 11,000 inhabitants and are excluded from the analysis.

### Implications for metapopulation disease models

After analyzing the stability of mobility patterns in both space and time, we evaluated how the spatiotemporal scale of human mobility affects our ability to effectively model metapopulation dynamics of disease. To address this, we integrate the connectivity network into a spatially explicit metapopulation model, with the goal of simulating the national spread of the initial wave of SARS-CoV-2 in the US. To understand the role of the geographic scale of mobility on disease dynamics, we integrate networks into the disease model that always at the US county level, but are homogenized at different spatial scales to represent missing information at different scales. To investigate the influence of temporal scale on model effectiveness, we inform the model with either a time-evolving connectivity network or a static connectivity network representing mobility from March, 2020 (without loss of generalizability given the temporal stability of the network we discuss above). In all cases, we measured goodness of fit by comparing the model predicted time of arrival of disease in a county to the observed time of arrival of the disease.

In Figure 4A, we demonstrate that a metapopulation disease model informed by a county-level intercounty connectivity network is highly predictive of observed early COVID-19 spatial diffusion. Relative to an Erdős–Rényi random intercounty connectivity network, the empirical mobility network has a stronger goodness of fit throughout the early phase of the pandemic. This emphasizes the crucial role of human mobility in the spatial spread of the initial SARS-CoV-2 wave and underscores the necessity of accessing mobility data for constructing more reliable models. Additionally, we find that the spatial invasion predicted by a static mobility network is highly consistent with the prediction from a time-varying mobility network, suggesting that static mobility data is sufficient to accurately reproduce epidemic spatial heterogeneity.

**Figure 4:**
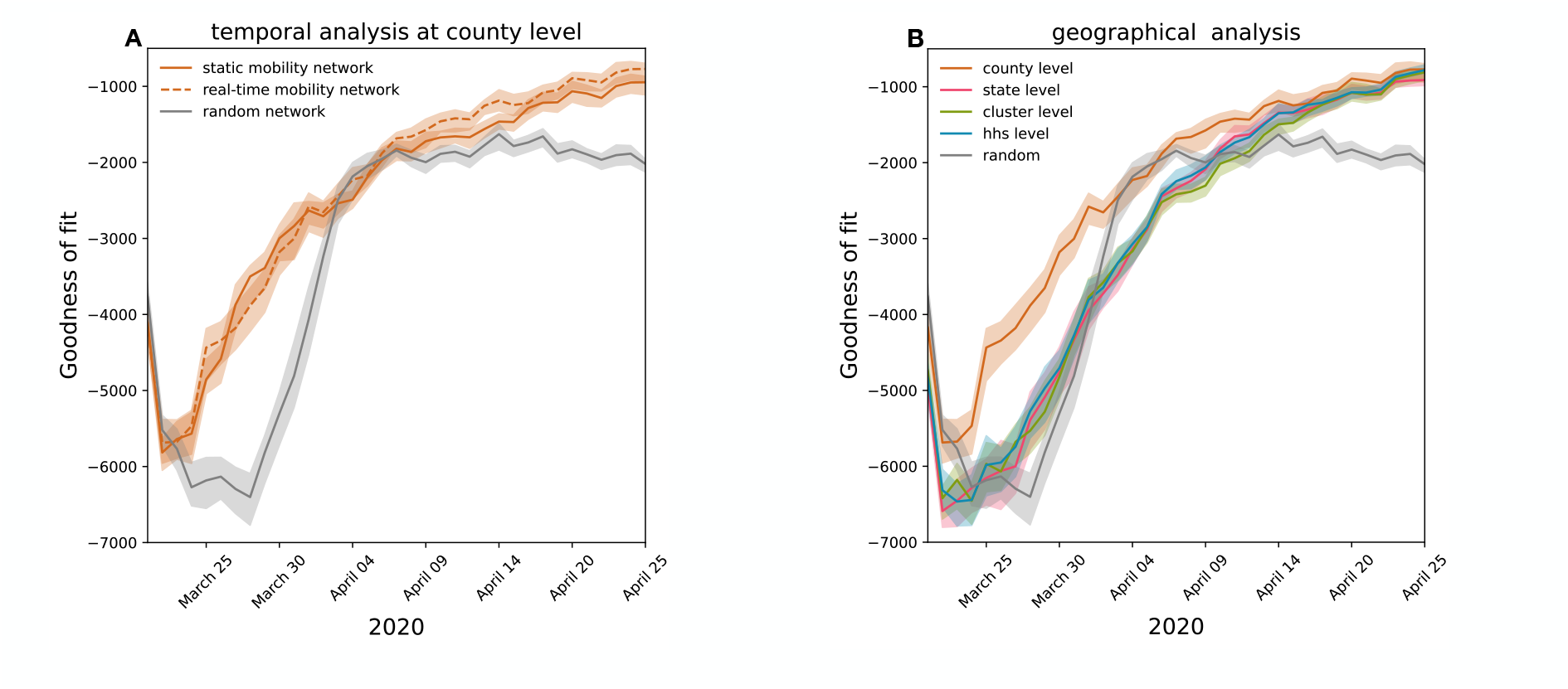
Implications of temporal and spatial scale of mobility data for prediction with metapopulation disease models. (A) The goodness of fit (median and 95% confidence interval) for time of arrivals for metapopulation models at a county level, informed by a time-evolving intercounty connectivity network, a static intercounty connectivity network, and a random intercounty network. (B) Solid lines show the goodness of the fit (median and 95% confidence interval) for metapopulation models informed with state-, cluster-, and HHS level mobility data.

In Figure 4B, we demonstrate the impact of the spatial scale of metapopulation structures on predictions of spatial diffusion. We compared the ability of county-level mobility data to predict spatial invasion, relative to mobility data at three spatial scales: US HHS regions, US states, and mobility data-based clusters (as defined in Figure 3). When comparing spatial scales, we find that all scenarios perform very similiarly to the random network. These results suggest that the state, cluster and HHS region-level mobility data lack the necessary granularity and fail to provide adequate insights into the diversity of mobility patterns.

## Discussion

Since the onset of the COVID-19 pandemic, mobile phone data has played a crucial role in addressing the public health crisis [27, 38, 35, 32, 20, 26, 34, 22, 43, 40, 29, 21, 19, 18]. During this period, numerous network operators and private enterprises have made considerable efforts to swiftly share their data within the confines of legal regulations. Consequently, researchers worldwide have embarked on working with this data, monitoring human behavior caused by containment measures and adaptive responses to the epidemic, and utilizing it to enhance epidemic models in order to increase their reliability. [22, 43, 40, 29, 26, 21, 20, 34].

While static mobility data have predominantly been analyzed and integrated into models over the past decades [4, 5, 6], the current accessibility to real-time human behavior data prompts an essential investigation into the optimal scenarios for utilizing this dynamic information versus relying solely on static representations of reality [30]. Equally important is the exploration of the characteristic mobility scale to comprehensively capture the intricate coupling between different locations, a consideration with potential implications for target control policies to reduce epidemic activity, and for improving epidemic model forecasting. Furthermore, numerous researchers have emphasized the pressing necessity to implement standardized strategies that facilitate rapid data access while upholding stringent data privacy measures [12].

To answer this gap in the literature, in this study, we investigate the spatial connectivity of US counties during the early phase of the COVID-19 pandemic using high-resolution real-time human mobility data obtained from mobile phone usage. Our findings reveal significant insights into the dynamics of human mobility and their implications for infectious disease modeling. In contrast to findings from other countries (e.g., France [22], India [23], Germany [24]), we observe that despite the implementation of local social distancing measures and lockdowns, intercounty connectivity remained largely unperturbed, leading to rapid geographic diffusion of SARS-CoV-2. Mobility patterns experience only marginal changes before and after the early-stage COVID-19 pandemic in March-April 2020. The most notable disruption occurred during the first lockdown period in April 2020, when mobility sharply declined. However, this reduction was short-lived, and mobility patterns quickly rebounded. Notably, even during periods of social distancing recommendations, the mobility network remains relatively stable. Assuming the lockdown represents the most extreme form of mobility disruption, the temporal stability findings suggest that global human mobility demonstrates resilience against short-term changes.

We also assess the spatial stability of the intercounty connectivity network by detecting spatial communities based on mobility patterns. Our results indicate that mobility-driven clusters align closely with state boundaries, reflecting the influence of administrative and geographical factors on human movement, accordingly with [8, 36]. These clusters exhibited remarkable stability over time, reinforcing the idea that spatial mobility patterns are deeply ingrained and relatively resistant to abrupt changes. The fact that mobility patterns are highly correlated with state boundaries suggests that state-level structures could be effective for designing target public health interventions based on travel reductions. Our findings underscore the importance of considering mobility patterns when designing interventions, resource allocation, and disease control strategies.

As shown in the context of COVID-19 pandemic in France [29], we also demonstrated that incorporating high-resolution human mobility data is crucial for accurately capturing the spatial spread of infectious diseases. Our findings indicate that county-level, daily mobility data offer the most accurate representation of the spatial spread of disease in the US. Notably, static county-level mobility data achieves similar model performance to real-time data, suggesting that an undisturbed representation of reality is adequate for reproducing spatial spread. More interestingly, our exploration of various spatial scales for metapopulation models underscores the significance of aligning the model’s structure with the inherent spatial scale of human mobility. While county-level mobility data yields the most accurate depiction, mobility data-based clusters, US states and HHS regions don’t capture the heterogeneity of the COVID-19 geographical diffusion.

While our study provides valuable insights, it is not without limitations. Our work focused on the early phase of the pandemic, during which response measures (e.g., social distancing, closures, (lack of) masking) were largely homogeneous in the US, and pharmaceutical measures (e.g., vaccination, antivrials) were not available; thus these findings are not generalizable to later stages. Furthermore, we assume homogeneity within US counties. Additionally, Safegraph mobility data, like all mobile phone datasets, exhibits sampling biases. On- going efforts to comprehend these biases are crucial for developing better correction methods [**coston“leveraging“2021**]. An independent analysis by Safegraph revealed the underrepresentation of older and non-white individuals in POI-specific analyses, though the panel is representative of race, education, and income [**coston“leveraging“2021**, 42].

While characterizing the key role of mobility in the spatial invasion of the COVID-19 pandemic in the US, our study sheds light on the global stability of human mobility patterns, and the relevant information needed to design a reliable predictive model. This result may be specific to countries such as the US in which mobility restrictions were not stringent, specified for intercounty mobility, nor enforced. Metapopulation models that incorporate accurate mobility data can provide valuable insights into disease dynamics and enhance our ability to predict and control the spread of future infectious disease outbreaks. Moreover, standardized data extraction and sharing we introduced might help facilitate the timelines associated with legal agreements for data sharing, which do not always align with the rapid spread of epidemics, thus diminishing the feasibility of timely responses to such outbreaks.

## Data Availability

mobility data were openly available to the public before the initiation of the study here:https://www.safegraph.com/
public health data are openly available here: https://covid.cdc.gov/covid-data-tracker/#datatracker-home
All data produced by data analysis and model simulations in the present work will soon be available on Github, here: https://github.com/GiuliaPullano/USA_first_wave_COVID_mobility.

https://www.safegraph.com/

https://covid.cdc.gov/covid-data-tracker/#datatracker-home

## Acknowledgments

The authors thank the teams at Safegraph/Advan Patterns for sharing mobility data.

