## Supplementary Information for "Characterizing US spatial connectivity: implications for geographical disease dynamics and metapopulation modeling"

### Data source

#### Mobile phone data

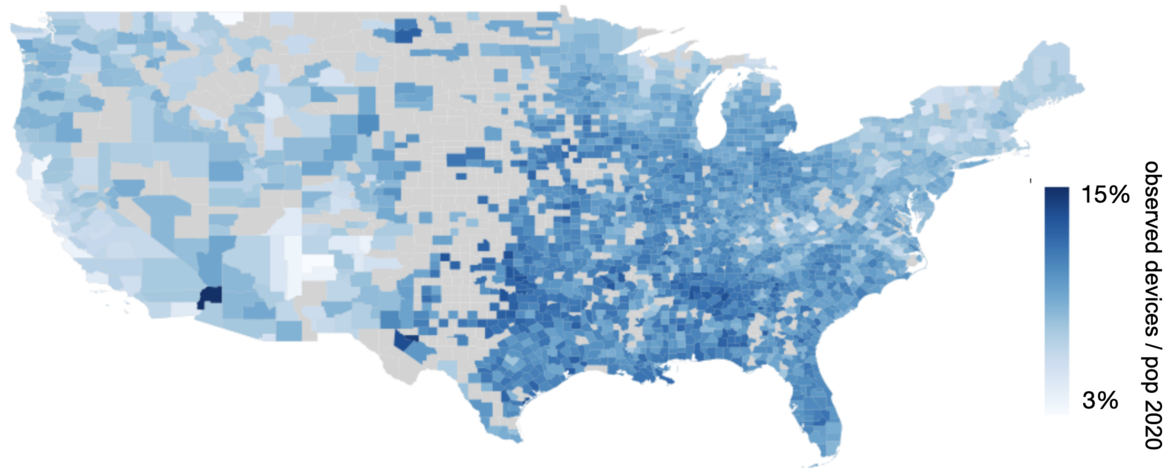


**Figure 1:** Mobile phone data coverage. Color-coded map of the ratio between observed mobile phone devices and population in 2020 by US county.

#### Comparing Neighbors patterns and social distancing dataset

SafeGraph's Neighborhood Patterns [1] and Social Distancing [2] dataset both contain footfall data aggregated by census block group (CBG) in the U.S. While the Social Distancing dataset does not account for any filtering procedure, Neighborhood Patterns (NP) does not report data unless at least 2 visitors are observed from census block groups. 48% of connections in SD are not present in the NP dataset. As the figure shows, the filtering process cut long-range connections, which are important from an epidemiological perspective. For this reason, we decided to use the daily SD dataset.

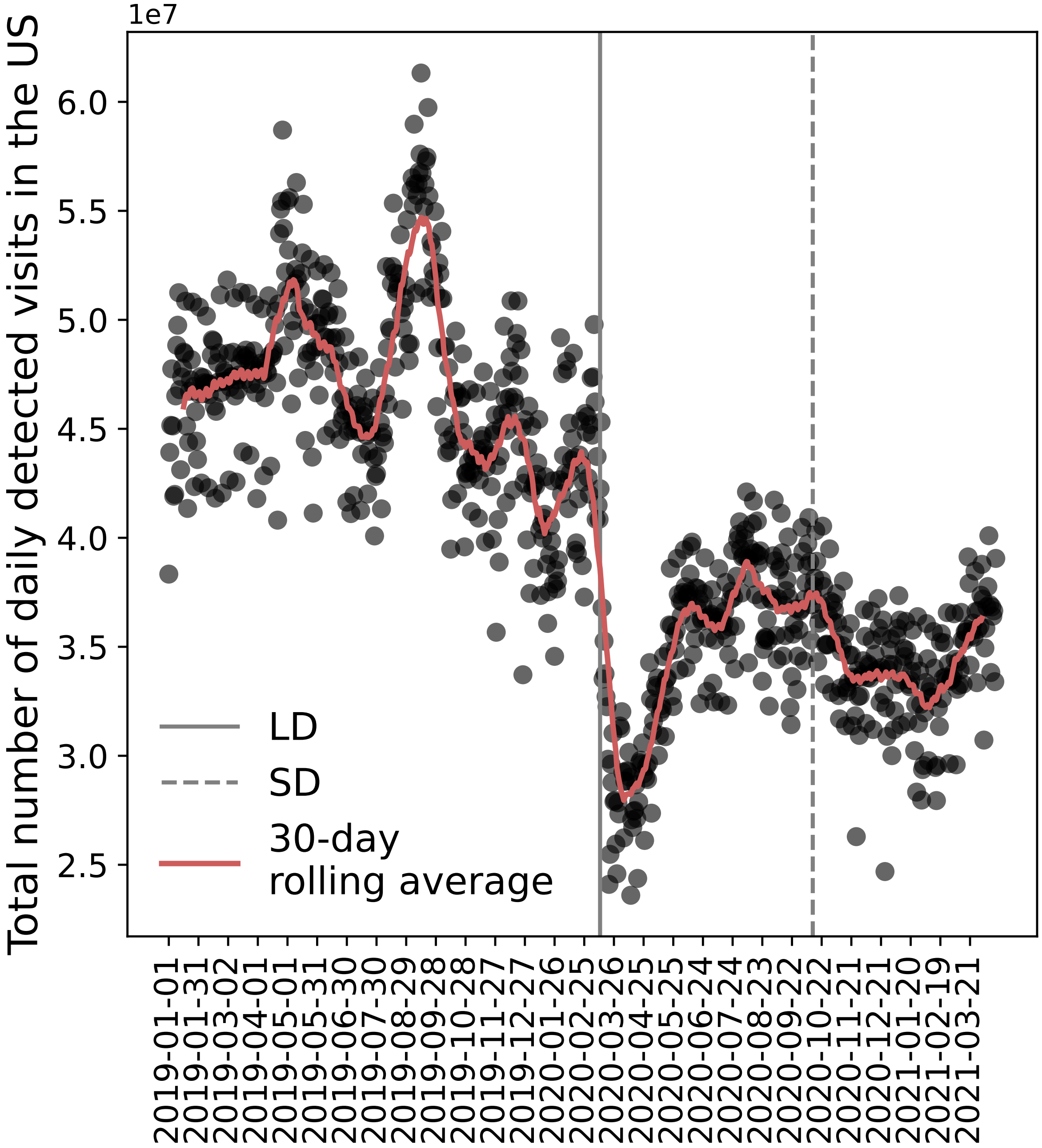


**Figure 2:** Detected mobility over time. Black dots show the daily total number of detected visits in the US provided by social distancing dataset. Solid brown lines show the 30-day rolling average. Vertical gray lines show Lockdowns (LD), and social distancing timelines (SD), respectively. Total number of devices revealed in any location summarized at the national level. The daily average is 40935499, with a 25% reduction at the end of March 2020 due to COVID-19 intervention policies.

### Additional results on the temporal stability

#### Characterization of intercounty connectivity network

##### Gravity Model

The gravity model is defined as follows:

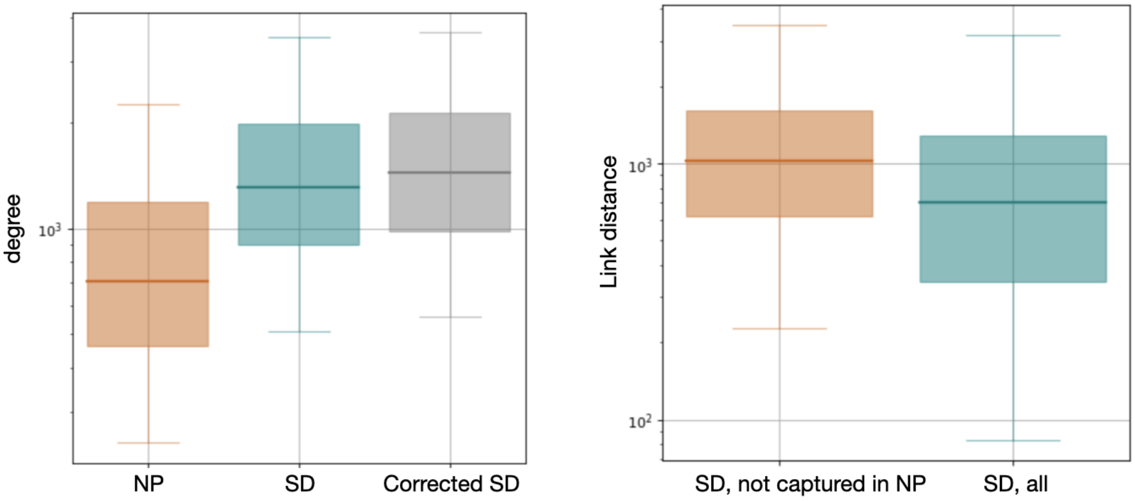


**Figure 3:** Comparison between SafeGraph's Social Distancing (SD) and Neighborhood Patterns (NP) dataset. Left plot: Box plots indicate the 95% reference range of the degree distribution in the intercounty connectivity network extracted using the NP dataset, the SD dataset, and the SD dataset corrected for heterogeneity in data coverage. Box plots indicate the 95% reference range of geographical distances between county connections in the SD network. The plot is broken down, accounting for all links and only the links not captured by the SD dataset.

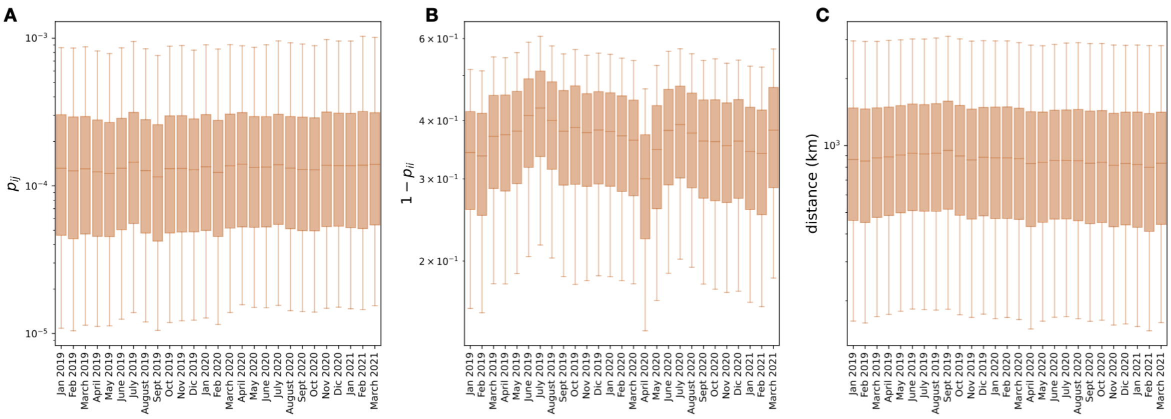


**Figure 4:** Temporal Stability of intercounty connectivity network. (A) The monthly connectivity between any pair of counties $i$, $j$, called coupling probability $p_{ij}$. Boxplots account for the 95th percentile of the distributions. (B) The monthly probability of going out of the residential counties (self-loop in the inter-county connectivity network). (C) Geographical distance between connected counties in the network. Boxplots account for the 95th percentile of the distributions.

$$p_{ij}^{gravity} = \left(P_i^{\gamma}\ P_j^{\theta}\right) \Big/ \left(a\ d_{ij}^{\delta}\right) \tag{1}$$

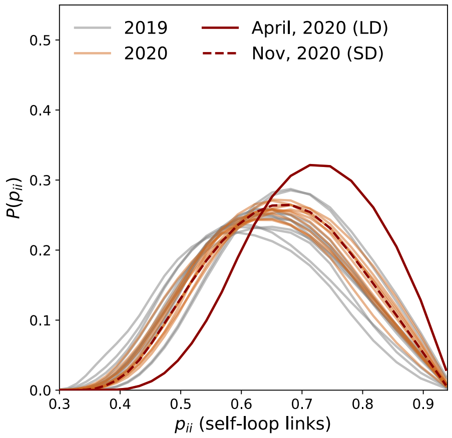


**Figure 5:** Probability of staying in the county of residence. The monthly probability of being in the residential counties (self-loop in the inter-county connectivity network). The probability of not moving during lockdowns in April, 2020 increased as expected.

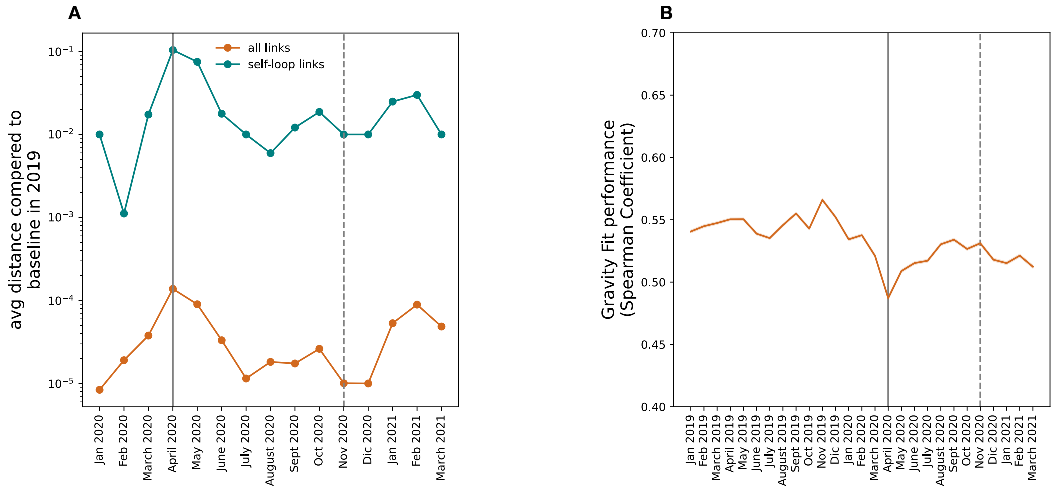


**Figure 6:** Connectivity network analysis over time. A) Average distance between the connetttivity network links in 2020, 2021 with the same month in a pre-pandemic period in 2019. B) Spearman Coefficient of the monthly Gravity Model. The Spearman coefficient over time illustrates the correlation between the observed intercounty connectivity network and the gravity network fitted using the observed intercounty connectivity network.

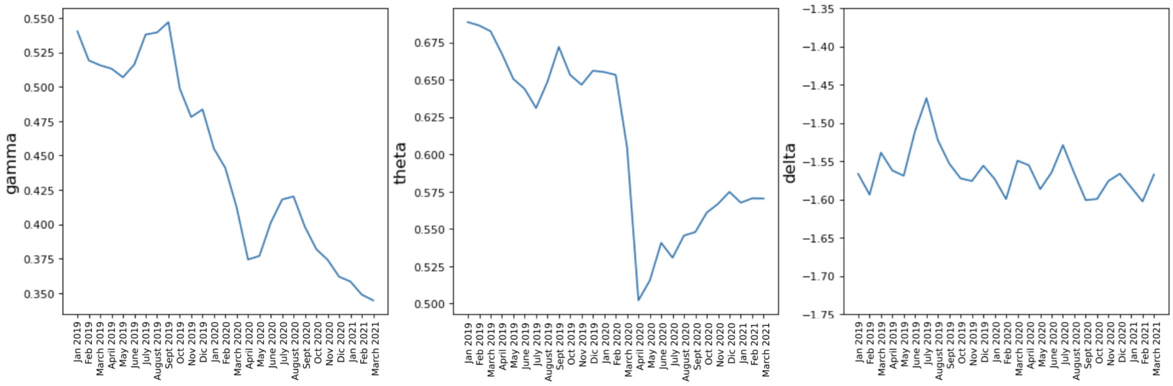


**Figure 7:** Fitted parameters of the monthly Gravity Model.

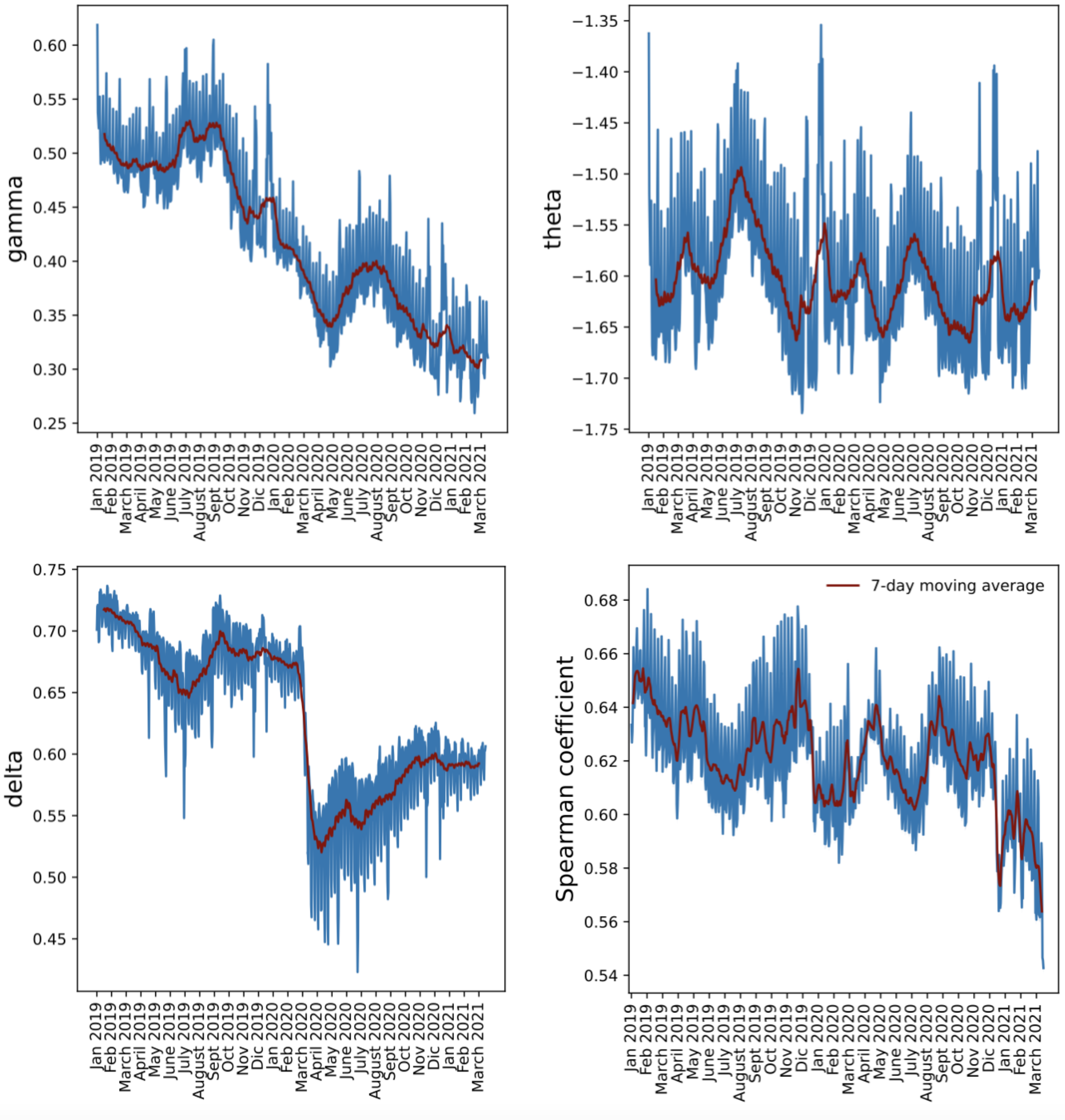


**Figure 8:** Fitted parameters and Spearman Coefficient of the daily Gravity Model. The Spearman coefficient over time illustrates the correlation between the observed intercounty connectivity network and the gravity network fitted using the observed intercounty connectivity network.

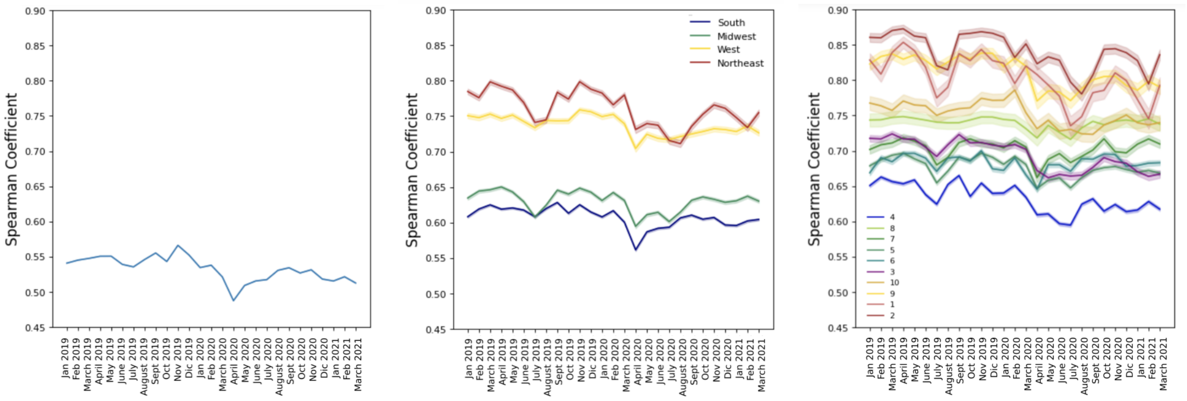


**Figure 9:** Spearman coefficient over time. The plots depict the performance of the fit across different spatial resolutions, including national, regional, and HHS office scales, respectively.

### Additional results on the spatial stability

To characterize spatial stability, we used INFOMAP algorithm. INFOMAP was developed particularly for mobility fluxes, and it uses the map equation and an information-theoretic approach, assuming that observed mobility flows are governed by a random walk process.

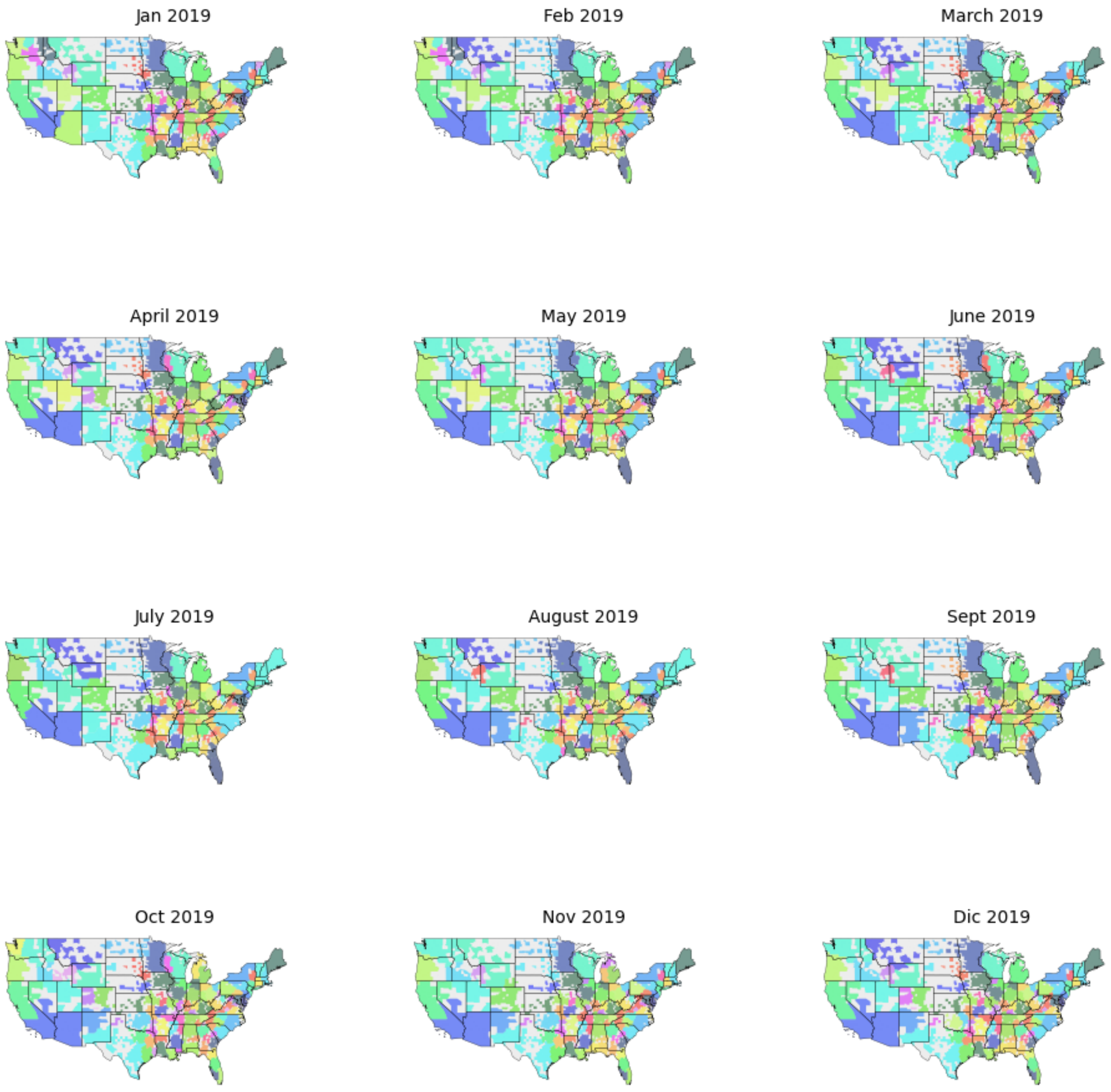


**Figure 10:** Monthly Infomap clusters in 2019.

#### Clustering analysis accounting for INFOMAP stochasticity

In order to assess the accuracy of the INFOMAP community detection algorithm we compute 25 simulations starting from the same seed. Then we compute the best partition of the system [**blondel2008fast**]. Besides, we compare the performance of INFOMAP with the Louvain clustering algorithm in Fig.12 finding a better characterization for the temporal evolution of the clusters with INFOMAP. In the case of Louvain, we sample 25 copies of the original network $G$, and for each copy, we draw for each edge a new weight, which is given from a random Poisson distribution with mean in the original weight. Then, we compute the best partition of the system.

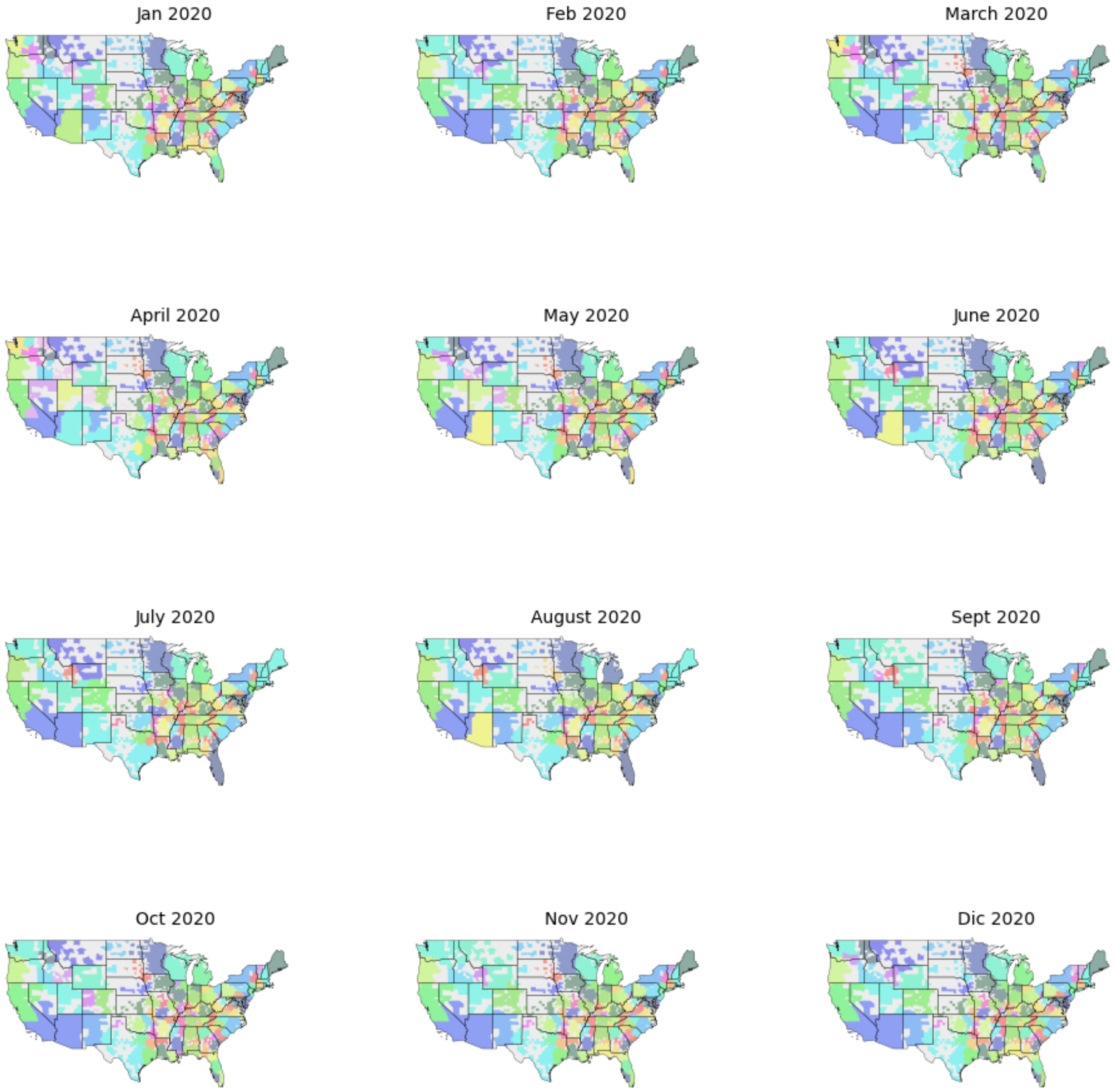


**Figure 11:** Monthly Infomap clusters in 2020.

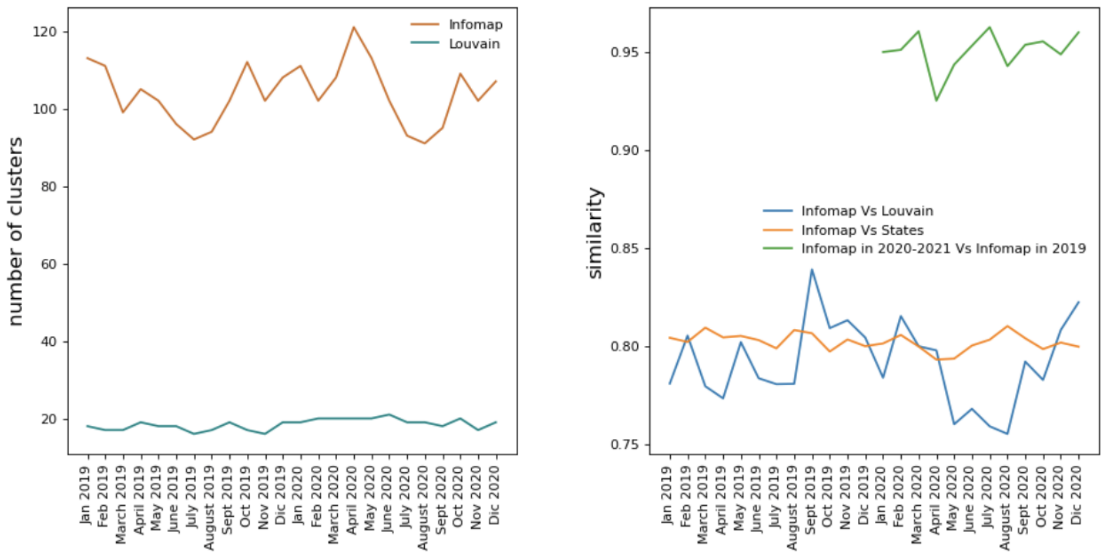


**Figure 12:** Cluster analysis. Left plot: number of clusters detected by Infomap and Louvain clustering algorithms. Right plot: monthly similarity between cluster repartitions.

#### Simulation details

The initiation of stochastic simulations is set for March 15, 2020. This date is deliberately chosen to focus exclusively on the national-scale invasion, disregarding any preceding international multi-seeding events as outlined in [3]. The model was initialized on reported cases on March 15, 2020, with necessary adjustments made to account for underreporting. Counties with reported cases before March, 15 are shown in Figure 1B. Underreporting was computed as defined in [4], and is shown in Figure 1C. For each simulation, the model outputs are the time of arrival of the first 10 infected cases in any county and the daily number of counties reached by the epidemic. A total of 60 stochastic simulations are performed using identical initial conditions. For the purpose of analyzing simulation outcomes, we compute the invasion probability $p_{i,inv}(t)$ for every county $i$ and time $t$. This probability denotes the likelihood of the epidemic reaching county $i$ by time $t$ [5]. It is calculated by dividing the number of runs in which county $i$ is impacted by the epidemic by the total number of runs conducted. In order to quantify the 95% confidence intervals (CIs) for the invasion probability, 60 runs were sampled with replacement 100 times, and relevant statistics were computed accordingly.

$$L(Nc_{obs}|\beta(T)) = \prod_{t=t0}^{T} P_{Poisson}(Nc_{obs}(t)|Nc_{pred}(t)) \tag{2}$$

### References


[1] *Neighborhood Patterns — SafeGraph. Link: https://docs.safegraph.com/docs/neighborhood-patterns.* en.

[2] *Social Distancing Metrics — SafeGraph. Link: https://docs.safegraph.com/docs/social-distancing-metrics.* en.

[3] Sen Pei, Sasikiran Kandula, and Jeffrey Shaman. "Differential effects of intervention timing on COVID-19 spread in the United States". eng. In: *Science Advances* 6.49 (Dec. 2020), eabd6370. ISSN: 2375-2548. DOI: 10.1126/sciadv.abd6370.

[4] *Using a delay-adjusted case fatality ratio to estimate under-reporting.* en. Mar. 2020.

[5] Michele Tizzoni et al. "On the Use of Human Mobility Proxies for Modeling Epidemics". en. In: *PLOS Computational Biology* 10.7 (July 2014), e1003716. ISSN: 1553-7358. DOI: 10.1371/journal.pcbi.1003716.
